# State-Level Gestational Diabetes Prevalence Estimates from Three Data Sources, 2018

**DOI:** 10.1101/2023.10.30.23297796

**Authors:** Michele L.F. Bolduc, Carla I. Mercado, Yan Zhang, Elizabeth A. Lundeen, Nicole D. Ford, Kai McKeever Bullard, Denise C. Carty

## Abstract

We investigated 2018 gestational diabetes mellitus (GDM) prevalence estimates in three surveillance systems (National Vital Statistics System, State Inpatient Database, and Pregnancy Risk Assessment Monitoring Survey). We calculated state GDM prevalence for each system; a subset of data was analyzed for women 18-39 years old in 22 locations present in all three systems to observe dataset-specific demographics and GDM prevalence using comparable categories. GDM prevalence estimates varied widely by data system and within the data subset despite comparable demographics. Understanding the differences between GDM surveillance data systems can help researchers better identify people and places at higher risk of GDM.

## Objective

We investigated differences in 2018 gestational diabetes mellitus (GDM) prevalence estimates using three surveillance data systems: National Vital Statistics System (NVSS), State Inpatient Database (SID), and Pregnancy Risk Assessment Monitoring System (PRAMS) data. GDM, an elevation of blood glucose concentrations initially detected during pregnancy, is associated with pregnancy complications and adverse outcomes such as perinatal mortality, macrosomia, and neonatal hypoglycemia (Yang et al. 2006). Women with GDM and their children are at higher risk of type 2 diabetes later in life (Bellamy et al. 2009, Dabelea et al. 2008). Some groups (e.g. older mothers, Asian mothers) are at higher risk of GDM and its complications (Li et al. 2020, Shah et al. 2021). Accurate GDM prevalence estimates are important to identify populations at higher risk; however, GDM prevalence estimates vary depending on the data source used. This paper presents state-level GDM prevalence estimates using three data systems and describes differences in prevalence by analyzing a comparable subset of the population (women 18-39 years old in 22 locations available in all three data systems). An understanding of the differences between GDM surveillance data systems, including their strengths and limitations, can inform public health policy and resource allocation.

## Methods

GDM prevalence was estimated using 2018 data from three surveillance systems: Centers for Disease Control and Prevention’s (CDC) National Center for Health Statistics’ National Vital Statistics System (NVSS) birth certificate data; Agency for Healthcare Research and Quality’s Healthcare Cost and Utilization Project State Inpatient Databases (SID) hospital discharge data; and CDC’s Pregnancy Risk Assessment Monitoring System (PRAMS) survey data.

Birth certificates are a complete enumeration of US births and are compiled in the NVSS. Birth certificates indicate the presence of GDM based on medical records. We queried the NVSS using CDC Wonder (https://wonder.cdc.gov/) for US states and the District of Columbia (DC). State-level GDM prevalence was calculated as GDM-associated births divided by total live births, using only singletons or the first birth in a set to avoid overcounting women with multiple births.

The SID is an unweighted census of more than 95% of hospital discharge records at the state-level. Live births to women affected by GDM were identified using diagnosis-related group (DRG) and International Classification of Diseases, Tenth Revision, Clinical Modification (ICD-10-CM) codes.^1^ State-level prevalence was calculated for the 27 states and DC available to CDC researchers, using total live hospital births with GDM divided by total live hospital births.

PRAMS is population-based surveillance survey of a sample of women with live births. Participants are sampled from birth certificates, with each participating state sampling between 1,000 and 3,000 women with a recent live birth per year. The survey contains a question about whether the respondent had gestational diabetes during their most recent pregnancy; participants self-report their GDM status. State GDM prevalence for 40 states and DC with response rates >50% in 2018 was calculated as births with GDM divided by total births among PRAMS respondents.

For comparisons, we also analyzed a subset of comparable data categories and locations that appear in all three data systems: women aged 18-39 years old with live births in 21 states and DC. Analyses were performed using SAS (version 9.4; SAS Institute), accounting for complex sampling in PRAMS.

## Results

Using NVSS data for all states and DC, state GDM prevalence ranged from 3.8% (Mississippi) to 11.0% (Alaska) [Map 1a]. Using the locations available in SID, GDM prevalence ranged from 5.4% (Mississippi) to 13.2% (Alaska) [Map 1b]. In locations with data in PRAMS, prevalence ranged from 4.5% in DC (95% CI: 2.5–6.4%) to 13.8% in Alaska (95% CI: 11.4–16.2%) [Map 1c].

**Map 1:**
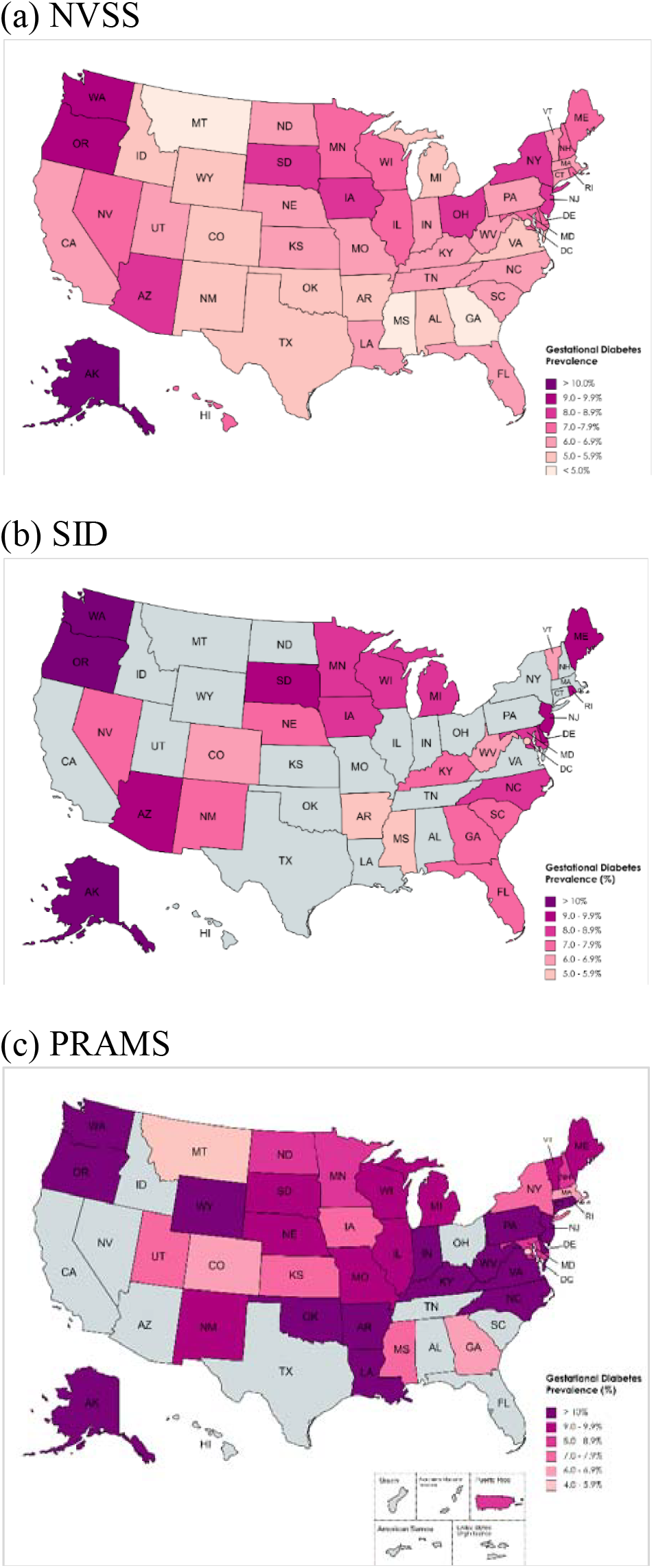
State-level gestational diabetes mellitus (GDM) prevalence estimates for 2018 as indicated by National Vital Statistics System (NVSS) (a), State Inpatient Database (SID) (b), and Pregnancy Risk Assessment Monitoring System (PRAMS) data (c). Greyed-out states do not have data available for analysis. Maps created at mapchart.net.

State GDM prevalence estimates varied widely by data system [Figure 1]. Among states with data in all three systems, West Virginia had the largest range in prevalence (6.1 – 11.7%). The smallest ranges in prevalence were in Colorado (5.3 – 6.4%) and Iowa (7.6 – 8.7%)

**Figure 1:**
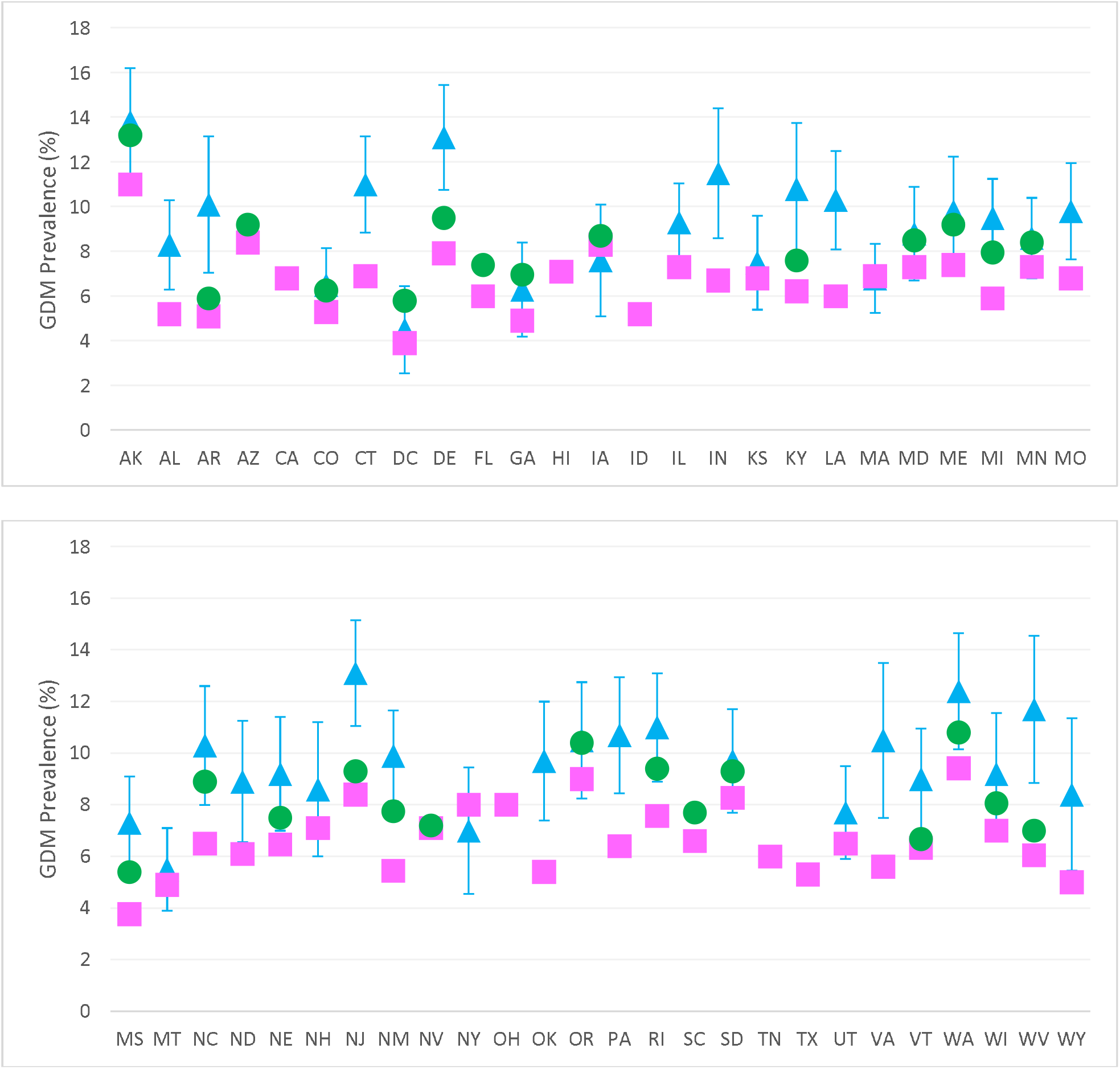
State-level GDM prevalence estimates for 2018 from National Vital Statistics System (pink square), State Inpatient Database (green circle), and Pregnancy Risk Assessment Monitoring System (PRAMS) (blue triangle). Blue error bars indicate 95% CIs for PRAMS estimates.

To understand how much of the differences in GDM prevalence across the data systems were related to differences in the populations included, we examined demographic differences between participants in a subset of comparable data (women 18-39 years old with live births in all data systems) for 21 states and the District of Columbia. Participant demographics varied only slightly by data system [Table 1]. NVSS contained a lower proportion of births to non-Hispanic (NH) mothers identifying with Another Race or more than one race then the other two systems; SID contained a lower proportion of births to Hispanic and NH Asian/Pacific Islander mothers; and PRAMS had a higher proportion of births to NH Black, NH White, and NH mothers of another race or more than one race. However, missingness of race and ethnicity was higher in SID (3.0%) and PRAMS (2.0%) than in NVSS (0.7%). Other demographics were similar across the data systems. Despite the similar populations in each system in our subsets, dataset-wide GDM still varied across the three data systems: 6.6% (NVSS), 8.0% (SID), and 9.0% (PRAMS; [95% CI: 8.5–9.6%].

**Table 1:** Demographic Characteristics for Women 18 – 39 Years Old Who Had a Live Birth in 22 Locations^a^ Represented in 3 Data Systems, 2018.

| Variable<sup>b</sup> | National Vital Statistics System | State Inpatient Database | Pregnancy Risk Assessment Monitoring System |
| --- | --- | --- | --- |
| Total Births | 1,049,098 | 990,115 | 1,008,398 |
| *Maternal Age* |  |  |  |
| 18–24 years | 249,765<br>(23.8%) | 235,252<br>(23.8%) | 234,529<br>(23.3%)<br>[95% CI = 22.4 – 24.2%] |
| 25–29 years | 323,385<br>(30.8%) | 304,924<br>(30.8%) | 308,994<br>(30.6%)<br>[29.7 – 31.6%] |
| 30–34 years | 317,465<br>(30.3%) | 300,173<br>(30.3%) | 310,396<br>(30.8%)<br>[29.8 – 31.7%] |
| 35–39 years | 158,483<br>(15.1%) | 149,766<br>(15.1%) | 154,480<br>(15.3%)<br>[14.6 – 16.1%] |
| Unknown | 0<br>(0.0%) | 0<br>(0.0%) | 0<br>(0.0%) |
| *Race/Hispanic Ethnicity<sup>c</sup>* |  |  |  |
| Hispanic | 157,586<br>(15.0%) | 123,568<br>(12.5%) | 142,471<br>(14.4%)<br>[13.8 – 15.0%] |
| Non-Hispanic | 884,007<br>(84.3%) | 836,509<br>(84.5%) | 845,486<br>(85.6%)<br>[85.0 – 86.2%] |
| American Indian, Alaska Native, Native American | 11,741<br>(1.1%) | 12,151<br>(1.2%) | 10,954<br>(1.3%)<br>[1.1 – 1.5%] |
| Asian or Pacific Islander | 59,384<br>(5.7%) | 49,317<br>(5.0%) | 49,543<br>(5.9%)<br>[5.5 – 6.3%] |
| Black | 177,144<br>(16.9%) | 170,748<br>(17.2%) | 160,670<br>(19.0%)<br>[18.1 – 19.9%] |
| White | 611,425<br>(58.3%) | 573,091<br>(57.9%) | 588,168<br>(69.6%)<br>[68.6 – 70.5%] |
| Another race/Multiple race | 24,313<br>(2.3%) | 31,202<br>(3.2%) | 36,151<br>(4.3%)<br>[3.9 – 4.7%] |
| Missing race or ethnicity | 7,505<br>(0.7%) | 30,038<br>(3.0%) | 20,441<br>(2.0%) |
| *Location* |  |  |  |
| Metro/Urban | 849,437<br>(81.0%) | 801,881<br>(81.0%) | 813,912<br>(80.7%)<br>[80.0 – 81.5%] |
| Nonmetro/Rural | 199,661<br>(19.0%) | 187,906<br>(19.0%) | 194,240<br>(19.3%)<br>[18.5 – 20.0%] |
| Missing | 0<br>(0.0%) | 328<br>(0.0%) | 246<br>(0.0%) |
| *Gestational Diabetes* |  |  |  |
| GDM prevalence | 68,962<br>(6.6%) | 79,328<br>(8.0%) | 89,995<br>(9.0%)<br>[8.5 – 9.6%] |
| Missing GDM | 1,149<br>(0.1%) | 0<br>(0.0%) | 11,182<br>(1.1%) |

<sup>a</sup> The locations included are AK, AR, CO, DC, DE, GA, IA, KY, MD, MI, MN, MS, NC, NJ, NM, OR, RI, SD, VT, WA, WI, and WV. ME and NE are also available in all three data systems for 2018; however, in SID, ME is missing age data and NE is missing race data, so these two states were excluded from the table.
<sup>b</sup> SID totals calculated as crude percentages using unweighted totals across the 22 locations. PRAMS were calculated using a cohort, showing weighted frequency, and weighted percentages in parentheses.
<sup>c</sup> Due to demographic limitations in the SID, women were categorized by ethnicity, and non-Hispanic women were further categorized by single-race and more than one race. SID also combines Asian and Pacific Islander women in one group. PRAMS data for Other Non-White and Mixed Race were combined for the Another Race/Multiple Race category.

## Discussion

We document variation in state-level GDM prevalence estimates by three unique data systems, and these variations exist even when controlling for similar population demographics. GDM prevalence estimates are influenced by the strengths and limitations of data systems, as well as screening practices that vary across facilities.

NVSS data provide a complete enumeration of live births in US states and territories and contain detailed demographic information. However, NVSS may underreport GDM; studies have documented a low sensitivity relative to medical records (46%–75.7%) (Gregory et al. 2019, Dietz et al. 2015, Devlin et al. 2009). Further studies are needed to understand how to improve documentation of GDM on birth certificates.

SID data have several strengths, including that they encompass more than 95% of US community hospital discharges; however, SID is derived from claims data, which are subject to coding errors and reflect only delivery hospitalizations. While NVSS indicates that <2% of births nationwide occurred outside of a hospital in 2018, some states have higher rates of non-hospital births, such as Alaska (7%). GDM pregnancies are at high-risk of adverse outcomes and may require hospital-based management during delivery. This may skew SID toward higher rates of GDM than would be seen in other birth settings. South Dakota, Rhode Island, West Virginia, and DC had more hospital deliveries identified in the SID than live births in birth certificate data, for unknown reasons. Finally, SID has less granular race and ethnicity data relative to the other two data systems used for this project, limiting our results to Hispanic and non-Hispanic single race categories.

PRAMS offers detailed survey data on health before, during, and after a live birth, connected to rich demographic data in birth certificates. However, survey response rates vary by site and have been decreasing over time; six states participating in 2018 did not meet the 50% response rate criteria. In addition, PRAMS is subject to biases associated with self-reported data (DeSisto et al. 2014, Dietz et al. 2014). A 2014 validation study of PRAMS data found moderate sensitivity and excellent specificity, but poor positive predictive values for self-reported GDM compared to medical records, indicating that self-report of GDM may be a valid information source (Dietz et al. 2014).

The US Preventative Services Task Force currently recommends screening for gestational diabetes in asymptomatic pregnant persons at 24 weeks of gestation or after. However, recommendations for screening protocols (one-step or two-step, Carpenter and Coustan or NDDG diagnostic cutoff values) vary by organization (USPSTF 2021). This lack of a universal standard for diagnosis of GDM may translate to a variable prevalence of GDM across medical facilities.

GDM screening combined with high-quality surveillance data can inform public health policy and resource allocation to prevent, identify, and manage this potentially serious pregnancy complication. Ideal GDM surveillance data could better identify the disproportionate burden of GDM across geography, race, ethnicity, and other demographic factors. While all three data systems explored here have strengths and weaknesses for GDM surveillance, birth certificates have the greatest potential for representativeness due to their enumeration of all births with detailed demographic information; however, improved surveillance, such as through hospital-based quality improvement initiatives, is needed to increase the accuracy of the data. All three data systems provide a useful source of information for prevention and management of GDM. Each could also be used by public health practitioners to increase availability of prevention programs to groups disproportionately impacted by GDM, such as the National Diabetes Prevention Program for which women with a history of GDM are eligible.

## Data Availability

NVSS data used for this project are publicly available via CDC Wonder (https://wonder.cdc.gov/natality.html). Access to HCUP and PRAMS datasets must be formally requested by researchers.

https://hcup-us.ahrq.gov/db/state/siddbdocumentation.jsp

https://www.cdc.gov/prams/prams-data/researchers.htm

## Acknowledgements

The views expressed in this manuscript do not necessarily reflect the official views or policies of the Centers for Disease Control and Prevention. Our work received no grant funding or separate financial support. No copyrighted materials were used in this article.

## Footnotes

1 If Diagnosis-Related Group (DRG) version is 35, then DRG codes 767-768 or 774-775 for vaginal delivery and 765-766 for C-section were used; if DRG version is 36, then DRG codes 768 or 796-798 or 805-807 for vaginal delivery and DRG codes 783-788 for C-section were used. Live births were identified using International Classification of Diseases, Tenth Revision, Clinical Modification (ICD-10-CM) codes delivery with at least one live birth (Z37.0, Z37.2, Z37.3, Z37.5, Z37.6). Births with documented GDM were identified by ICD-10 codes O24.4 or O99.81 listed anywhere on the discharge record. ICD-10 codes for pre-pregnancy diabetes were excluded (E08, E09, E10, E11, E13, O24.0, O24.1, O24.3, O24.8, or O24.9).

## Notes

### Competing Interest Statement

The authors have declared no competing interest.

